# Transmission and epidemiological characteristics of Severe Acute Respiratory Syndrome Coronavirus 2 (SARS-CoV-2) infected Pneumonia (COVID-19): preliminary evidence obtained in comparison with 2003-SARS

**DOI:** 10.1101/2020.01.30.20019836

**Authors:** Rongqiang Zhang, Hui Liu, Fengying Li, Bei Zhang, Qiling Liu, Xiangwen Li, Limei Luo

## Abstract

**Objectives:** Latest epidemic data of Severe Acute Respiratory Syndrome Coronavirus 2 (SARS-CoV-2) infected Pneumonia (COVID-19) was collected and a detailed statistical analysis was carried out to make comparison with 2003-SARS in order to provide scientific reference for the prevention and control of COVID-19.

**Methods:** The information of COVID-19 and 2003-SARS from websites of NHCPRC and the World Health Organization was collected, and then the transmission dynamics of the two kinds of infectious diseases were analyzed. The information of 853 confirmed COVID-19 patients obtained from the website of health committees of 18 provinces. A descriptive epidemiological analysis method was employed to carefully analyze the epidemic characteristics. Subsequently, the COVID-19 epidemic data in Wuhan and other inland regions of China was analyzed separately and compared. A multivariate function model was constructed based on the confirmed COVID-19 case data.

**Results:** The growth rate of new cases and deaths of COVID-19 were significantly faster than those of 2003-SARS. The number of confirmed cases in Wuhan and other inland areas both showed increasing trends. 853 confirmed COVID-19 cases aged 1 months to 94 years and the average age was (45.05 ± 17.22) years. The gender ratio (M: F) was 1.12: 1.

**Conclusions:** The fatality rate of COVID-19 is lower than that of 2003-SARS and the cure rate is higher. The age of COVID-19 patients is mainly concentrated in the 30-50 years old (60.61%). The harm of the first-generation COVID-19 patients is higher than that of secondary cases.

---

On December 8^th^, 2019, the National Health Commission of the People’s Republic of China (NHCPRC) of the People’s Republic of China received a case report of an unexplained pneumonia patient. Subsequently, NHCPRC appointed an expert group to arrive in Wuhan to guide the epidemic management. On January 7, the pathogen of this unexplained infected pneumonia was identified as a novel coronavirus (2019-nCoV) ^[1]^, which was renamed as Severe Acute Respiratory Syndrome Coronavirus 2 (SARS-CoV-2) on February 12^th^, 2020. SARS-CoV-2 could causes Coronavirus disease 2019 (COVID-19), also known as SARS-CoV-2 infected Pneumonia. On February 7^th^, 2020, a total of 31774 cases of COVID-19 patients were diagnosed and reported in 31 provinces (autonomous regions, municipalities) and Xinjiang Production and Construction Corps (XPCC)^[2]^. At the same time, China’s Hong Kong Special Administrative Region, Taiwan, Macau and Japan, the United States, Vietnam, Singapore, Nepal, France, Canada, South Korea, Thailand, Malaysia, etc, all have COVID-19 reports. The rapid spread of COVID-19 has brought a huge impact on the health of Chinese residents, economic development, and even social stability.

Within the last week, Chinese governments had taken a series of comprehensive measures to prevent and control the Novel Coronavirus (COVID-19) Pneumonia (COVID-19), such as closing the traffic of Wuhan to minimize the spread of confirmed cases, doctors and nurses across the country and around the world to assist Wuhan. Scientists have also actively researched and explored the etiology, genomics, transmission dynamic of COVID-19 and made due contributions to the prevention and control of the spread of COVID-19 infection. However, from the perspective of the epidemic of COVID-19, its number of confirmed cases is still increasing rapidly, and the number of deaths has also increased significantly. Actively exploring and studying the transmission characteristics and epidemiological characteristics of COVID-19 will greatly help to curb its spread. It has been reported that the epidemiological and clinical characteristics of COVID-19 show some similarities to Severe Acute Respiratory Syndrome in 2003 (2003-SARS) ^[3-4]^. Whether the effective control of 2003-SARS can provide direction for the prevention and control of COVID-19, there is still no reported evidence. In the present study, we collected relevant latest epidemic data of COVID-19 and a detailed statistical analysis was carried out and the results were compared with 2003-SARS in order to explore and provide scientific reference for the prevention and control of COVID-19.

## Materials and Methods

### Diagnosis and Definition of COVID-19 Patients

A confirmed COVID-19 patients was defined as a case with respiratory specimens that tested positive for the SARS-CoV-2 by at least one of the following three methods: isolation of SARS-CoV-2 or at least two positive results by real-time reverse-transcription-polymerase-chain-reaction (RT-PCR) assay for SARS-CoV-2 or a genetic sequence that matches SARS-CoV-2 ^[5]^.

### Epidemic Trends of COVID-19

The data of confirmed COVID-19 cases from January 10^th^, 2020 to February 7^th^, 2020 and 2003-SARS cases from March 26^th^ to May 26^th^, 2003 in China’s inland regions (data from websites of NHCPRC and the World Health Organization) was collected and then the epidemic trends of the both infectious diseases were analyzed to understand the transmission dynamics of them.

### Epidemiological Characteristics of COVID-19

The basic information of COVID-19 patients reported in inland areas of China (Beijing, Shaanxi Province, Sichuan Province, etc.) was collected, including age, gender, and contact history (consist of having been to Hubei or living in Hubei, contacting confirmed cases, contacting people who were from Hubei and unclear). The data was collected from the website of the health and health committees of the above provinces. A descriptive epidemiological analysis method was employed to carefully analyze the epidemiological characteristics of COVID-19. However, only several provincial administrations released the baseline information of COVID-19 confirmed patients, so the number of confirmed cases with detailed baseline information was far less than the total. Finally, 853 COVID-19 confirmed patients from 18 provinces, whose baseline information was available, were included into our analysis.

### Comparison of COVID-19 in Wuhan and Regions except Wuhan

COVID-19 originated in Wuhan, China. In the early stage of COVID-19 outbreak, there may be cases of underreporting or concealment in Wuhan, which may make it difficult to objectively reflect the true epidemic characteristics of COVID-19. Therefore, the COVID-19 epidemic data in Wuhan and other inland regions of China was analyzed separately and compared to obtain more objective COVID-19 epidemic knowledge.

## Results

### Overall epidemic trends of COVID-19

As of 24:00 on February 7^th^, NHCPRC has received a total of 31774 confirmed COVID-19 cases in in 31 provinces (autonomous regions, municipalities) and Xinjiang Production and Construction Corps (XPCC), 6401 severe cases, 722 deaths, and 2050 discharged patients. There were 27657 suspected cases. The spatial trend of the COVID-19 outbreak was showed in Figure 1. The temporal trend of the cumulative number of COVID-19 and 2003-SARS patients was showed in Figure 2.

**Figure 1.**
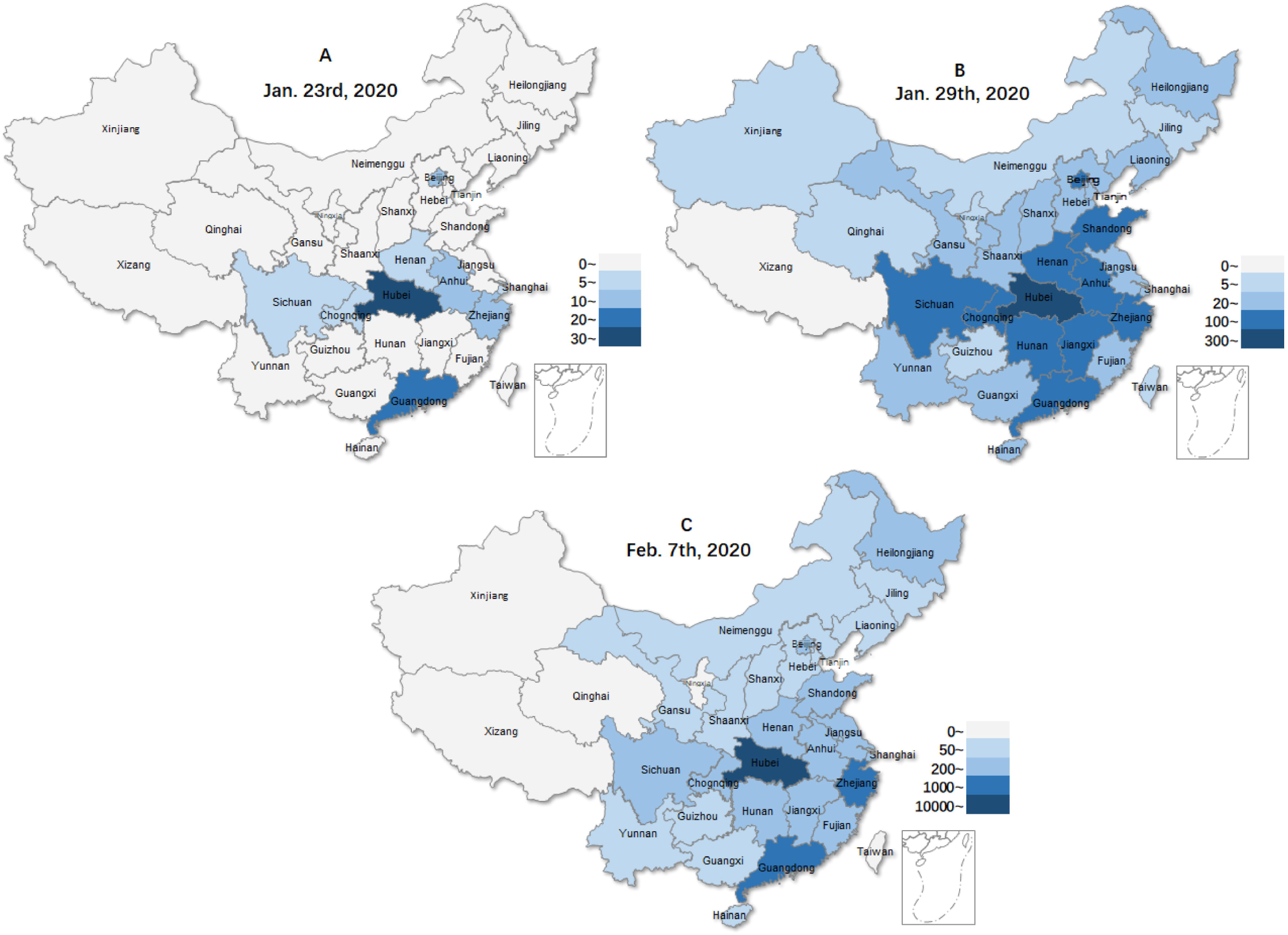
The spatial trend of the COVID-19 outbreak.

**Figure 2.**
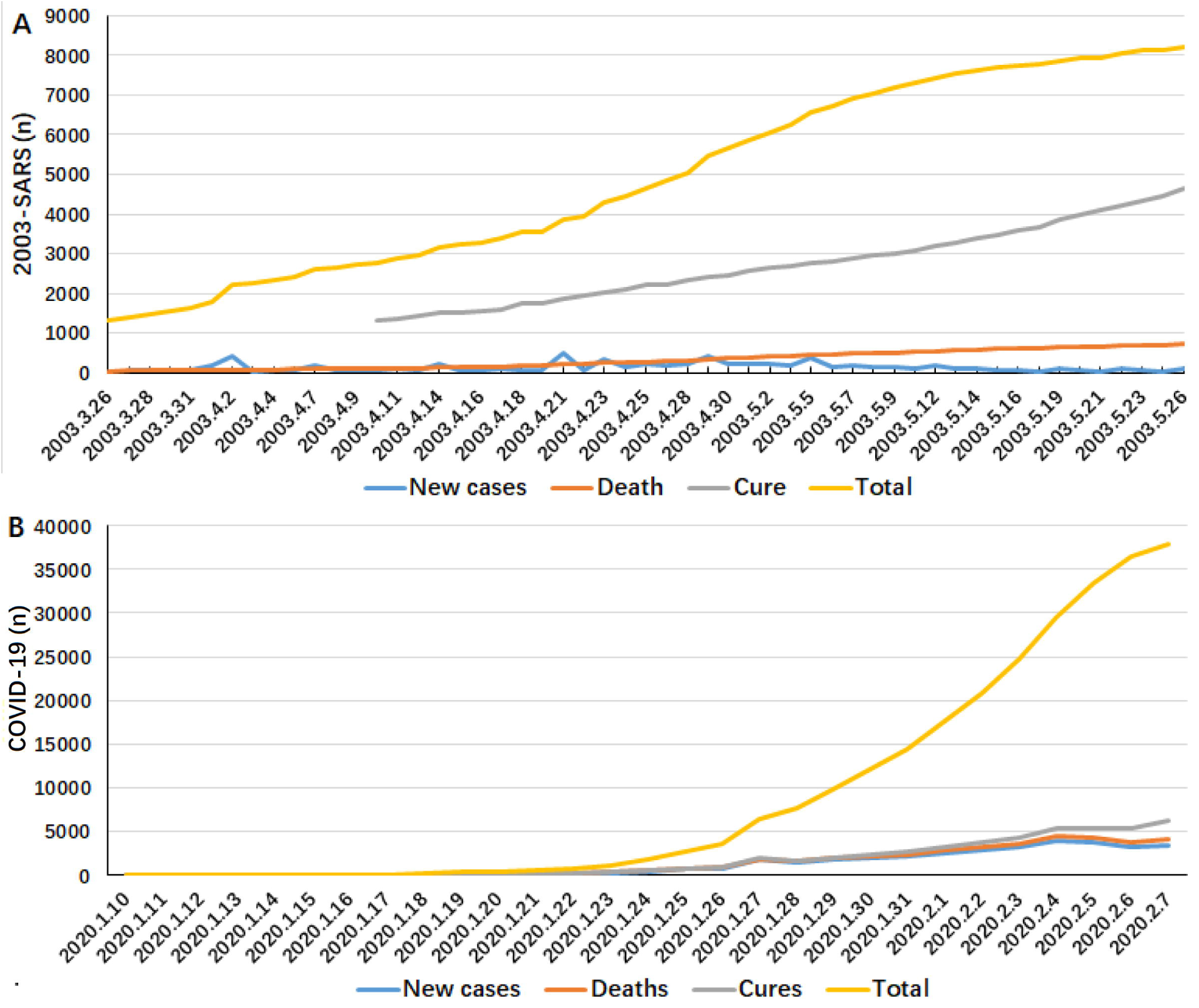
Comparison of the epidemics between COVID-19 and 2003-SARS.

### Epidemic trends and characteristics of COVID-19 and 2003-SARS

As shown in Figure 3, the growth rate of new confirmed cases and deaths of COVID-19 were significantly faster than those of 2003-SARS in the first month after the outbreaks of both infection diseases, while the number of cured patients was significantly less than that of 2003-SARS, suggesting that COVID-19 showed a faster transmission and a longer treatment.

**Figure 3.**
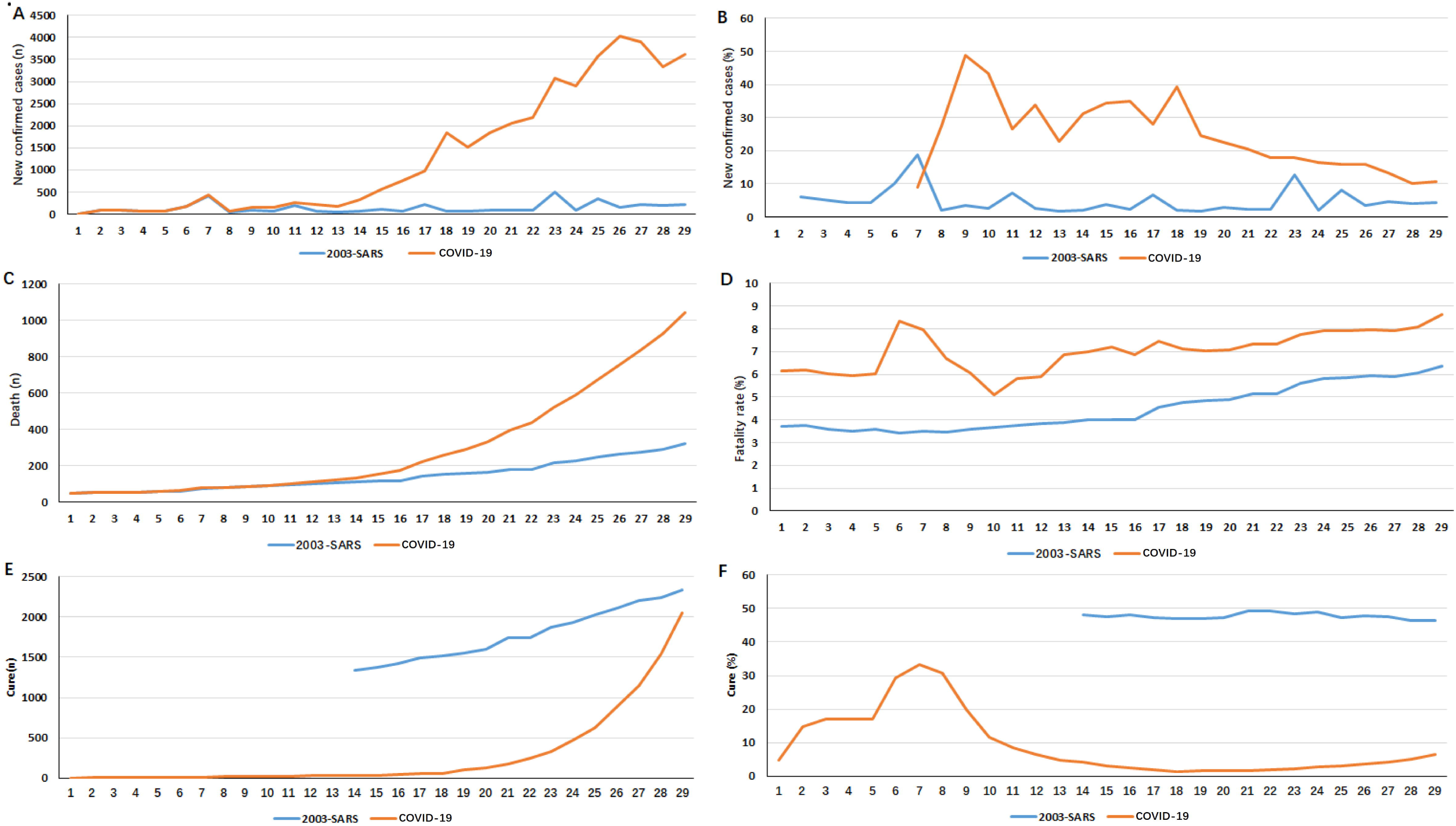
Comparison of trends in confirmed cases and fatality rate of COVID-19 between Wuhan and regions except Wuhan (not include the data of Hongkong, Macao, and Taiwan; Days for COVID-19 were January 10^th^ from February 7^th^, 2020; Days for 2003-SARS were March 26^th^ from April 13^th^, 2003).

### Trends in Confirmed Cases and Fatality Rate of COVID-19 of Wuhan and Regions except Wuhan of China’s Inland

Both the numbers of confirmed cases in Wuhan and other inland areas showed increasing trends. From January 24^th^, 2020, the number of confirmed cases in areas other than Wuhan in inland China began to exceed Wuhan (Figure 4A); the fatality rate in Wuhan (1.52% -6.64%) was significantly higher than that of inland areas of China except for Wuhan (0% -2.64%) (Figure 4B).

**Figure 4.**
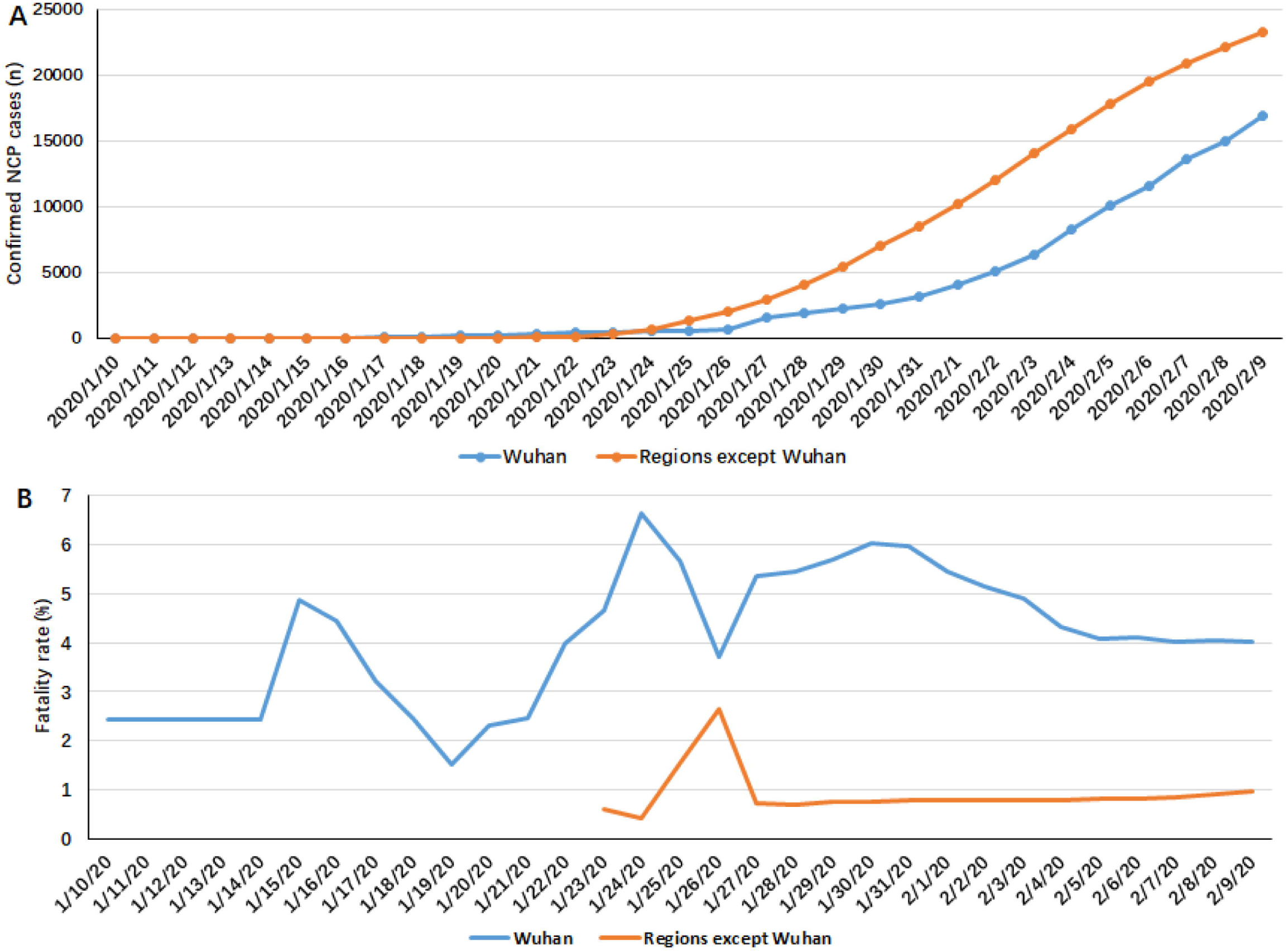
Trends in confirmed cases and fatality rate of COVID-19 of Wuhan and regions except Wuhan of China’s inland.

### Epidemiological Characteristics of COVID-19 patients

As of 17:00 on February 7^th^, 2020, the basic information of 853 confirmed COVID-19 cases was collected on the official websites of 18 provincial and municipal health committees. 853 confirmed COVID-19 cases were aged 1 months to 94 years and the average age was (45.05 ± 17.22) years; 853 confirmed COVID-19 cases included 450 males (52.75%) and 403 females (47.25%), the gender ratio (M: F) was 1.12: 1 (Figure 5, Table 1).

**Table 1.** Characteristics of age distribution of 853 COVID-19 patients n(%)

| Age group (Y) | Male | Female | Total |
| --- | --- | --- | --- |
| 0- | 7(26.92) | 19(73.08) | 26 |
| 10- | 18(62.07) | 12(37.93) | 29 |
| 20- | 48(54.55) | 40(45.45) | 88 |
| 30- | 109(55.90) | 86(44.10) | 195 |
| 40- | 101(59.76) | 69(40.24) | 169 |
| 50- | 75(49.02) | 78(50.98) | 153 |
| 60- | 56(42.42) | 76(57.58) | 132 |
| 70- | 26(57.78) | 19(42.22) | 45 |
| 80- | 9(64.29) | 5(35.71) | 14 |
| 90- | 1(50.00) | 1(50.00) | 2 |

**Figure 5.**
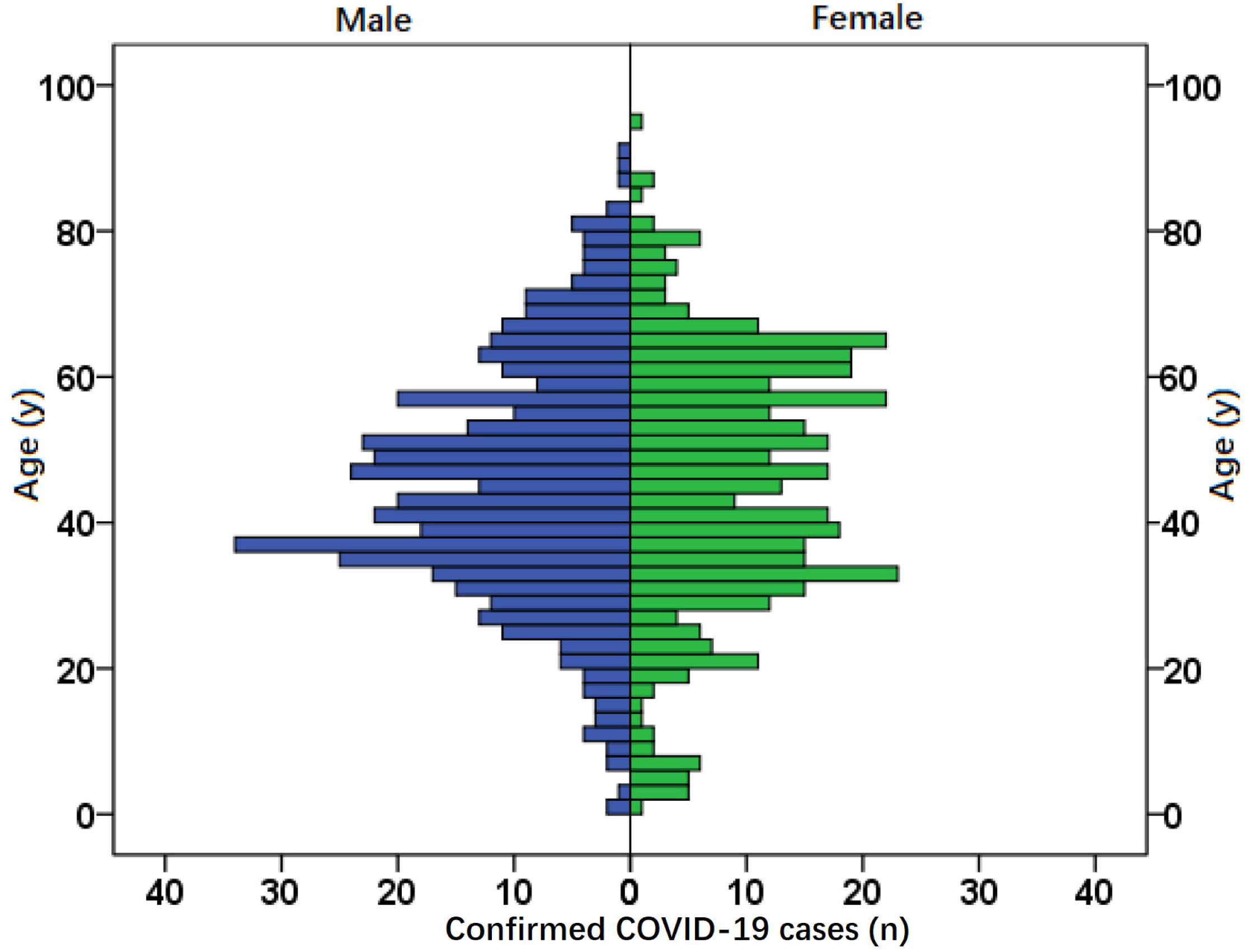
Population pyramid plot of 853 confirmed COVID-19 patients.

In the 853 confirmed COVID-19 patients, 199 cases reported their contact history, in which 172 patients (86.43%) had a history of having been to Hubei or living in Hubei, 19 patients (9.55%) had a history of contacting confirmed cases, 8 patients (4.02%) had a history of contacting people who were from Hubei (Figure 6).

**Figure 6.**
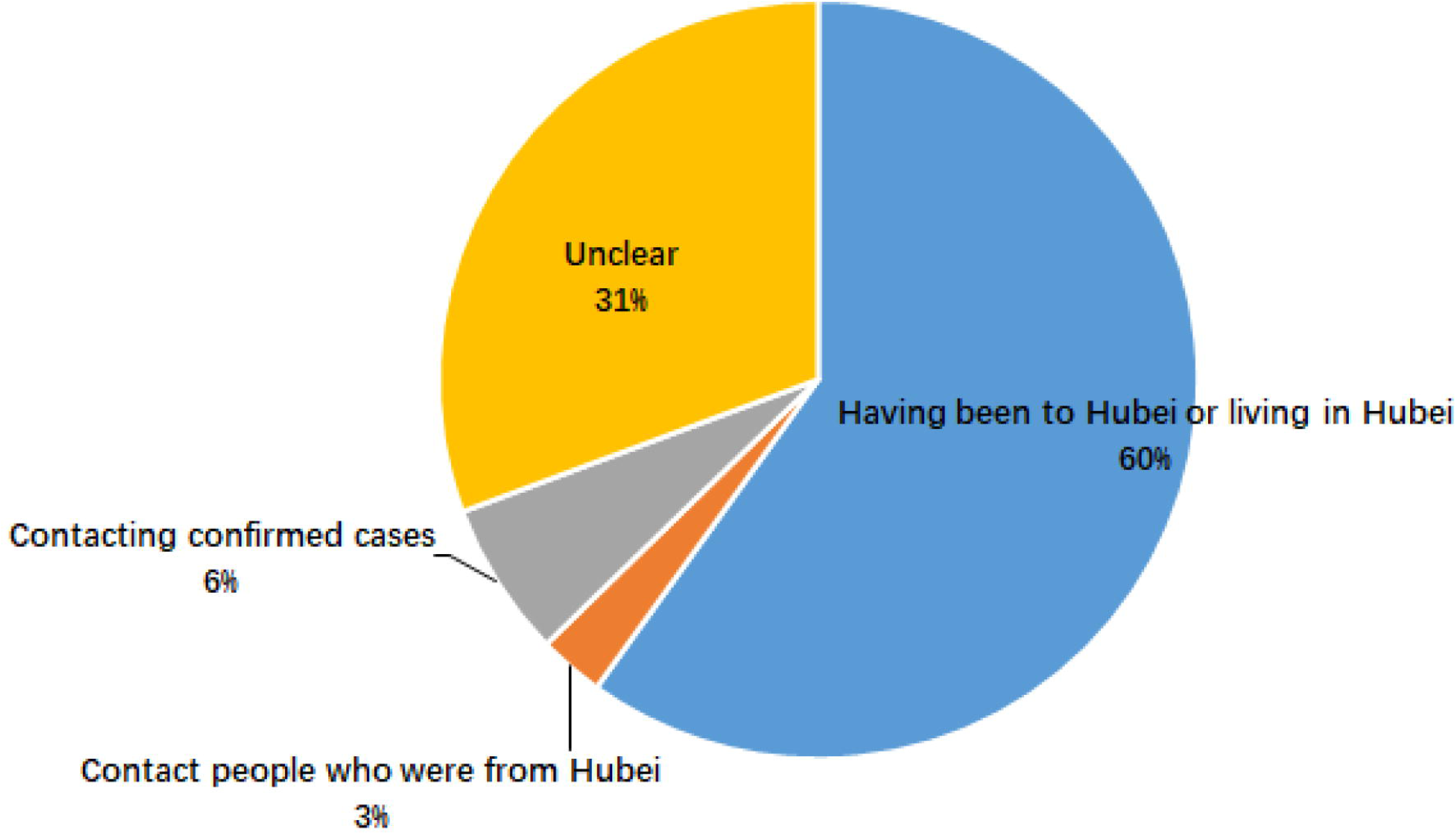
Contact history of 199 confirmed COVID-19 patients.

## Discussion

In 2002, a case of SARS appeared in Guangdong Province, China, and then spread to Southeast Asia and all of the world ^[6]^. The SARS outbreak comprehensively tested that the public health system of China was very vulnerable in front of the prevention and control of infectious diseases. Subsequently, the Chinese government implemented a series of reforms in the public health field, such as reorganization of the CDC, training public health professionals, and establishing the disease information system covering the whole country, the establishment of a complete reporting system for infectious diseases and an excellent mechanism for handling public health emergencies, etc. After a development for 17 years, a complete public health system has been established in China and great progress has been made in handling public health emergencies such as infectious disease epidemics.

After the outbreak of COVID-19 in 2020, the Chinese government adopted comprehensive measures to handle its spread. The people from China and the world also provided generous assistance to Wuhan, including actively treating the confirmed patients, donated medical supplies such as masks, etc., which have been played important roles in effectively curbing the spread of COVID-19. The main process of COVID-19 in China was shown in Figure 7. Meanwhile, governments across China were also actively adopting strict measures. The appropriate performance of the Chinese government and local governments at all levels to deal with COVID-19 was due to the experience accumulated in the SARS disposal process and the great development of public health system in the past 17 years.

**Figure 7.**
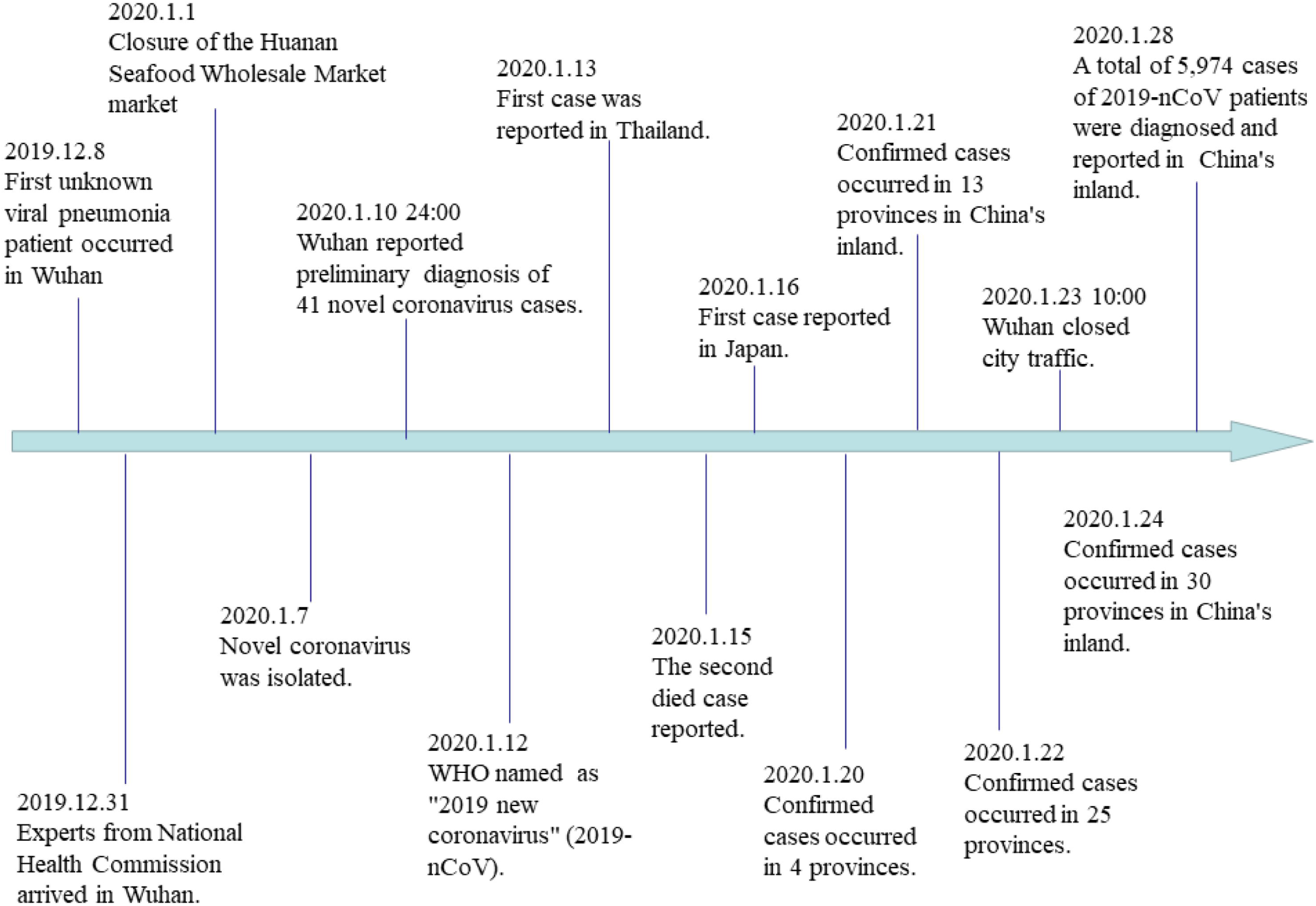
The main process of the early stages of COVID-19 outbreak in China.

The COVID-19 epidemic has brought a significant impact on the health and normal lives of the residents and the social and economic development of China. In addition, it was catching up with the Chinese Lunar New Year, and the above adverse effects are even more prominent. However, until now, the etiology, epidemiology, and pathogenesis of COVID-19 are unclear, which has made it difficult to effectively contain the epidemic. Under this background, the latest data of COVID-19 epidemic was collected to conduct a detailed analysis, and compared with the relevant characteristics of 2013-SARS. The aim of our present study is to make positive contributions to help stop the COVID-19 epidemic and to restore China’s normal social order as soon as possible.

Although many details such as the source of the virus and the ability to spread among people are still unknown, an increasing number of cases that have confirmed the human-to-human transmission of the virus. Studies have shown that the interpersonal transmission ability of COVID-19 corresponds to SARS, but its doubling time (6 to 7 days) is shorter than that of SARS (about 9 days) ^[1, 7]^, and the passage interval of COVID-19 is also relatively short. The second generation of COVID-19 cases unrelated to Wuhan has gradually appeared, which has also contributed to a sharp increase in short-term confirmed cases, resulting in a serious situation of COVID-19 prevention and control, and thus it is now very difficult to predict the epidemic trend.

Age and comorbidities (such as diabetes or heart disease) are independent predictors of poor SARS-CoV and MERS-CoV outcomes. The spread of SARS-CoV and MERS-CoV occurred to a large extent through super-transmission events ^[6-7]^. Currently, there are data suggesting that there may be super-transmission events in COVID-19 transmission, and the entire range of interpersonal transmission capabilities of COVID-19 remains unknown ^[8-9]^. In addition to the fragility of medical institutions in the face of the new coronavirus epidemic, the risk of infection complications among hospital populations is also greatly increased ^[10-11]^. What we need to do now is to continue to strengthen prevention and control measures, actively strengthen patient admission, diagnosis, and treatment.

Our results showed the following epidemiological characteristics related to COVID-19:

1. The overall epidemic of COVID-19 is similar to that of 2003-SARS, but the total number of confirmed cases is higher than that of 2003-SARS. The fatality rate is significantly lower than that of 2003-SARS in the same period, and the cure rate is significantly higher than that of 2003-SARS, which suggests that the prognosis for COVID-19 is good.
2. The age of COVID-19 patients is mainly concentrated in the 30-50 years old (60.61%), and the male are more susceptible than the female (M: F 1.12: 1). This may be related to the fact that the adults at this age group are more active in social activities than other age groups.
3. The results showed that that the fatality rate of COVID-19 in Wuhan (1.52% -6.64%) was significantly higher than that in other regions (0%-2.64%). Most of the confirmed cases in Wuhan were the First-generation infections, and the other regions may exist more secondary cases. The information above provided evidence that the harm of the first-generation COVID-19 cases is significantly higher than that of secondary cases (including second- and third-generation cases).

## Data Availability

All data generated or analyzed during this study are available from the authors upon reasonable request.

## Acknowledgments

The present study was funded by Research Project from Health Commission of Shaanxi Provincial Government (2018A017), Research Project from Education Department of Shaanxi Provincial Government (19JS015), Subject Innovation Team of Shaanxi University of Chinese Medicine (132041933).

## Author contributions

R. Z. and X. L. planned the study.

R. Z., H. L., F. L. B. Z., Q. L. and L. L. performed the data analysis, wrote and modified the manuscript.

R. Z. submitted the study.

